# Pharmacometric Generative Stochastic Modeling of Patient Reported Outcome Measures

**DOI:** 10.1101/2025.03.12.25323869

**Authors:** Kuteesa R. Bisaso, Karyaburo R. Kadada, Karungi S. Bisaso, Jackson K. Mukonzo, Ene I. Ette

## Abstract

**Background/Objectives:** Patient reported outcome measures (PROMs) capture the patient’s own perspective on their health, illness, and therapeutic effects on the illness. However, their analysis and interpretation is challenging due to their multidimensional nature, poor correlation with clinical and physiological outcomes, lack of a standardized interpretation, and discrete nature of the data. We describe a generative stochastic modeling approach and show that it improves the pharmacometric characterization of multi-item PROMS.

**Methods:** The Restricted Boltzmann Machine (RBM) modelling approach was described and used to model the relationship between efavirenz mid-dose concentrations,clinical variables (CD count and Viral load) and time varying patient reported neuropsychological impairment symptoms. The model was used to derive a variable importance ranking for all the PROM items, clinical variables, and drug concentrations.

**Results:** The model adequately characterizes the PROMs. Variable importance ranking reveals that mid-dose concentrations are not more predictive of post-baseline PROMs than clinical variables and baseline PROMs.

**Conclusions:** Generative stochastic modeling with RBMs adequately characterizes PROMS and their relationship to other variables and drug concentrations, is readily adaptable to the pharmacometric workflow, and is able to generate individual level disease progression trajectories using baseline variables.

## Introduction

Patient-reported outcome measures (PROMs) are questionnaires used to capture the status of a patient’s health or illness directly from the patient, devoid of any interpretation from the clinician or caregiver.^1^ They capture the thoughts, complaints, opinions, and other subtle outcomes that would otherwise be impossible for anyone other than the patient to describe. Moreover, it is imperative to incorporate the patient’s own perspective on the illness and the effect of therapy in regulatory decisions affecting quality of life.^2^ Data obtained from PROMs are used to assess disease outcomes and measure the benefits or risks of intervention for the patient. But in order to obtain a claim on a drug label, the PROM has not only to detect a change in disease status, but its analysis should be valid and reliable. Patient-reported outcome measures often produce numerical scores that are typically bounded, discrete, and skewed, which complicates the choice of statistical methods to use for their analysis. Moreover, one patient-reported outcome (PRO) scale can generate multiple outcomes, reported in different forms, based on different score types, sub-scales, or summary scores, potentially increasing the complexity of analysis. Traditional approaches to the analysis of patient-reported outcomes include logistic regression for binary and ordinal outcomes, linear mixed-effects, linear regression, and analysis of covariance for multivariable analysis.^3^

Pharmacometric approaches, which integrate pharmacological and biological mechanisms underlying a phenomenon of interest, are frequently applied to the analysis of PROMs. Unlike their statistical counterparts, pharmacometric approaches seek to estimate the exposure-response profile and use that to predict, via simulation, the effect of treatment over time in various unstudied situations.^4^ Some pharmacometric analyses of PRO data include repeated time-to-event analysis of event data, Poisson and negative binomial models of count data, logistic regression of binary data, proportional odds analysis of ordinal categorical data, Markov modeling of discrete (and sometimes continuous) time data, bounded integer analysis of composite scores and linear regression or linear mixed-effects modeling for composite scores.^5–7^ These methods analyze a single PROM or a composite of multiple items of a PROM. Despite the ease of computation and analysis, the use of composite scores results in item-level loss of information. A common approach is to estimate each scale separately, using Classical Test Theory or, more recently, Item Response Theory (IRT), which maximizes knowledge extraction by simultaneously modeling item-level data.^8,9^

Longitudinally, pharmacometric PRO models estimate disease progression trajectories of individual patients and incorporate the effect of treatment. Disease progression modeling uses mathematical functions to describe quantitatively the time course of disease progression. Disease progression models can be broadly categorized into three classes: empirical, semi-mechanistic, and systems biology. Of these, the empirical category is the simplest, is purely data-driven, and does not describe underlying biological processes. Empirical models provide mathematical frameworks for interpolation between observed data.^10^ They facilitate the use of PROMs to monitor the natural history of diseases and detect treatment effects.^11^ Various functional forms have been employed to empirically model disease progression using PROMs ranging from simple linear constant disease rate models to complex generalized logistic functions incorporating beta-distributed residual variability and covariate sub-models.^12,13^

However, a common challenge of analyzing PROMs using disease progression models is the potential for model misspecification arising from the violation of modeling assumptions, as each method has its own assumptions, estimation procedures, and estimands. The general assumptions when analyzing PROMs include the normality of residuals and linearity of the relationship between the outcome and independent variables or covariates. Their violation results in unreliable clinical estimates and predictions, leading to inaccurate, erratic, and non-robust decision-making. These challenges could be resolved by the use of an analysis method such as generative stochastic modeling that is capable of simultaneously modeling item-level data, covariates, and drug effects without making any relational, structural, or distributional assumptions.

Generative stochastic models are capable of learning complex patterns and new representations in data, constructing joint distributions of all variables, and generating new data instances that resemble the data on which they are trained.^14,15^ In addition, they are capable of inferring missing observations and unobserved patterns via semi-supervised learning and self-supervised learning, respectively. They can apply the learned dependencies to a wider range of functions from those studied.^16^ Generative models are building blocks of artificial intelligence.

The objective of this study is to demonstrate the adaptation of generative stochastic modeling to pharmacometric characterization and simulation of PROMs with an additional application to therapeutic drug monitoring (TDM).

## Methods

### The Generative Model

Generative models learn the complex statistical distribution of an input dataset with the goal of generating new data points that resemble the input dataset. Energy-based generative models define an energy function that produces low energy values when the combination of all inputs is compatible and higher energy values when they are not.^17^ They have deep connections to statistical physics whose purpose is to derive observable quantities from microscopic laws governing the constituents of a system^18^. The most prominent energy-based generative model is the restricted Boltzmann Machine (RBM).

Restricted Boltzmann Machines are energy-based probabilistic generative neural networks made up of one visible layer and one hidden layer, each with several nodes. All the nodes in the visible layer are connected to all the nodes in the hidden layer but no connections exist between nodes of the same layer. The empirical data is represented by the visible nodes, while the stochastic hidden nodes capture the structural features in the data and build arbitrary dependencies among the visible nodes into a joint distribution.^19^

Being energy based models, an energy function E(v,h) is defined and exploited to define the joint distribution between visible (v) and hidden (h) nodes, namely the Boltzmann distribution.

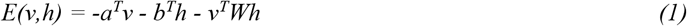

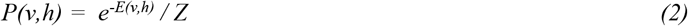

where Z is a partition function that makes the probabilities sum up to one and is defined as the sum of *e*^*-E(v,h)*^ over all possible pairs of visible layers and hidden layers, *W* are the parameters connecting the visible and hidden nodes, also referred to as the weights, while *a* and *b* are the baseline intercepts of the visible and hidden nodes, also referred to as biases. Since the nodes in each layer are mutually independent of other nodes in the same layer, the conditional probabilities of the hidden *(P(h*|*v))* and visible *(P(v*|*h))* nodes are defined as

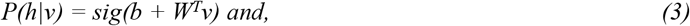

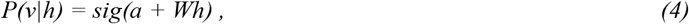

where *sig* is the sigmoid function.

Instead of taking the mean sigmoid output as the value of nodes, the conditional probabilites are used to sample the activations of the hidden and visible units generate to their respective states

The marginal probability assigned to nodes of the visible layer is obtained by summing over all hidden nodes by

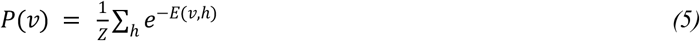

When marginalized over all possible configurations of hidden nodes, the expectancy or model distribution of any given configuration of visible nodes is measured by its free (visible) energy given by.

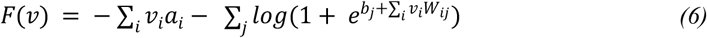

It is the energy that a single visible layer pattern would need to have in order to have the same probability as all of the configurations that contain *v*. ^19^

Being generative rather than discriminative neural networks, the loss function of RBMs is not straightforward or differentiable. Back-propagation cannot be used to compute the gradients of the loss function with respect to the model parameters. Instead, learning the RBM amounts to finding a statistical distribution such that data sampled from this distribution is statistically indistinguishable from the input data. Thus, the gradient of the log probability with respect to the weights is given by:

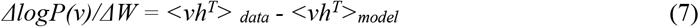

where the angle brackets denote the expectations of the distribution specified by the data and model subscripts. The expectation with respect to the model (*<vh*^*T*^*>*_*model*_) is intractable, and its estimation via Markov chain sampling is slow. Therefore, its estimation is performed through a Gibbs sampling method called contrastive divergence.^20^ Contrastive divergence (CD) is an approximation technique primarily used to train energy-based models, an example of which is the RBMS. The Gibbs chain is initialized with input data v_(0)_ and runs for k steps (usually k = 1 is sufficient but higher k values are better), yielding the sample v_(k)_. Each Gibbs step t consists of sampling h_(t)_ from p(h|v_(t)_), then sampling v_(t+1)_ from p(v|h_(t)_) using equations (3) and (4). See Figure 1.

**Figure 1.**
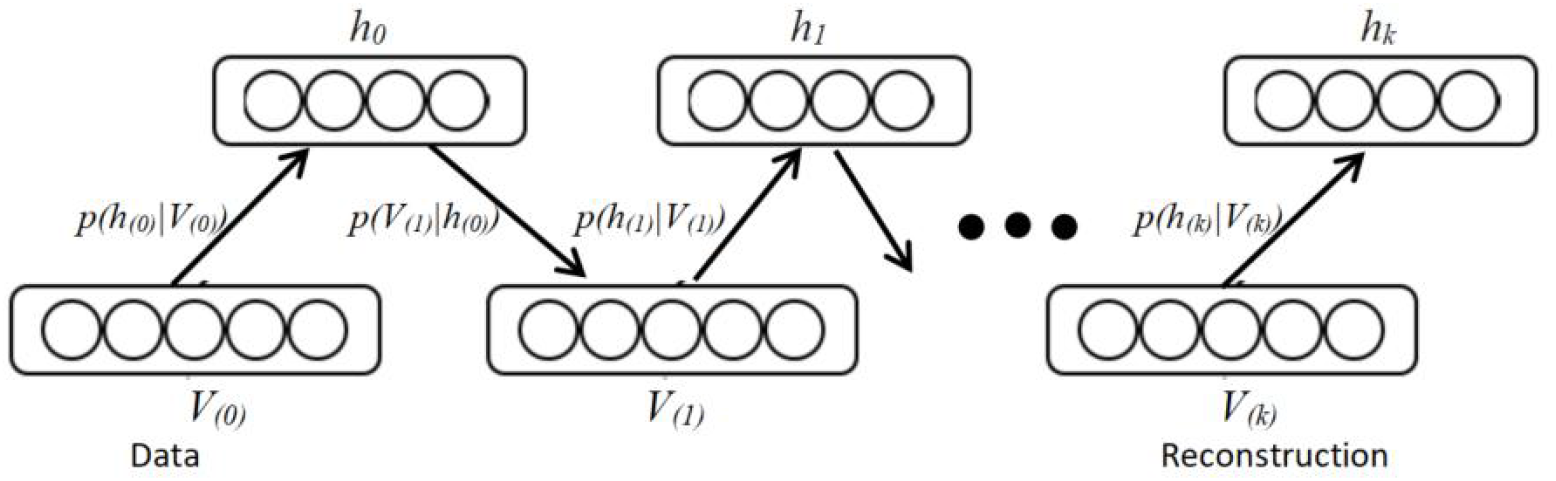
A diagram of K-steps Gibbs sampling of an RBM with 5 visible nodes and 4 hidden nodes.

The update in parameters is given by

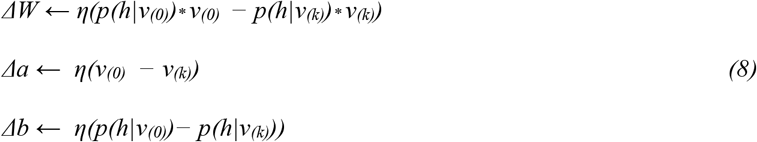

where Δ means change in the respective parameter and η is the learning rate.

Restricted Boltzmann machines were initially developed for binary visible and hidden nodes. There have been later modifications that handle real-valued visible nodes, including Gaussian, categorical, and count data by modification of the energy function to form the mixed variate RBM.^21^

When dealing with Gaussian visible nodes, the conditional probability of the visible nodes is given by:

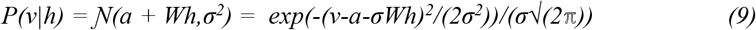

where *σ* is the standard deviation of the visible unit, and *𝒩* is the Gaussian function.

Likewise, the energy function is given by;

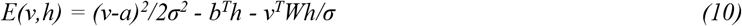

In addition, it was later revealed that noisy rectified linear hidden nodes (nReLu) were better at extracting complex features than the original binary nodes.^22^ The CD objective function, which initializes the visible units with the data for every training chain, has also been modified by using the persistent contrastive divergence (PCD) objective function which initializes the visible units with persistent samples from the previous iteration/training chain, thereby letting them run longer via Gibbs sampling. This allows the samples to better represent the model’s equilibrium distribution, reducing bias and improving convergence, especially for complex distributions‥^23^ Additional improvements involve the inclusion of the reconstruction error in the gradient approximation,^24^ penalizing the cost function by restricting the free energy of the training data by the addition of the free energy to the update rule in equation 8,^25^ and the use of adversarial training methods in combination with the maximum likelihood.^26^ The reconstruction error, free energy, and adversarial gradients are added to the CD gradient in equation 8 before it is used to update the parameters. Their contribution to the update rule is controlled by regularization parameters. All these improvements apply to the binary as well as mixed variate RBMs.

### Data

To demonstrate the modeling of multi-item PROMs with RBMs, a neuropsychological impairment (NPI) dataset reported by Bisaso et al and Mukonzo et al.^7,9,27^ was used. It comprised patient-reported outcomes from 196 HIV patients on 600 mg efavirenz-based antiretroviral therapy (ART) evaluated at baseline, days 14 and 84. A total of 4136 observations of seven symptoms (items) describing sleep disorders, hallucinations, and cognitive impairment; baseline CD4 count, baseline viral load, and steady-state mid-dose concentration (C_p,ss,(12 h)_) calculated from individual efavirenz pharmacokinetic empirical Bayes estimates (EBE) namely, clearance (Cl), absorption rate constant (K_a_), volume of distribution (V_d_), and bioavailability fraction (F1) as follows

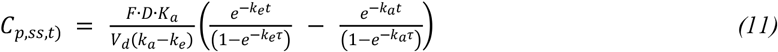

Where t = 12 hours and τ is the dosing interval (24 hours), k_e_ is the elimination rate constant calculated as Cl/V_d_. Efavirenz PK parameters used in this investigation were obtained from our previous work.^7^ The neuropsychological PROMs, namely, insomnia, vivid dreams, sleepwalking, audio hallucinations, visual hallucinations, tactile hallucinations, and Mini Mental State Examination (MMSE) score category at baseline and on days 14 and 84, were included as individual variables. Each patient provided a single row capturing all their data. The covariate data types were categorized as continuous, binary, or categorical. The steady-state mid-dose concentration, viral load, and CD4 count were continuous variables, while the PROMs, except MMSE, were binary variables, and the MMSE -an ordered categorical variable with three categories (normal = 0, moderate = 1, and severe = 2).

### Software

The analysis was done in R version 4.2.2. ^28^. No additional modeling R packages were used.

### Model Training

The continuous variables were transformed and scaled to the standard normal distribution, N(0,1). Binary variables were coded as 0 and 1, while the categorical variables were one-hot encoded. One-hot encoding is a method for converting categorical variables into a binary format for each category in the original variable to improve the model’s performance.^29^ Binary nodes were used for the hidden layer instead of nRELU units to avoid complication of the free energy function that is later used to monitor training and measure variable importance. The mixed variate RBM model was trained by minimizing the Boltzmann encoded adversarial persistent contrastive divergence (k = 20) cost function penalized by reconstruction error and free energy.^21,23–26^ Hyperparameters were selected for the model by grid-searching all possible combinations using 5-fold cross-validation in which the model with a selected combination of hyper-parameters was trained on four parts of the data and tested on the left-out fifth part of the dataset. The hyperparameters included learning rate (0.0001, 0.001), weight decay (0.0001, 0.001), number of hidden nodes (20, 50, 100, 200), regularizers of free energy in the cost function (0, 0.001, 0.01), and the contribution of the adversarial component of the training process (0.3, 0.7, 1). The batch size was arbitrarily set to 30 and the training momentum to 0.6. Each model was set to run for 1000 epochs. An epoch is a complete pass of a training dataset through the learning algorithm. After training, each model was tested by reconstructing the left out fifth part, and the reconstruction error as well as the free energy calculated. The process was repeated until each of the five parts had been used as the left-out fifth part for testing, and the average reconstruction error of the five parts was calculated for each combination of hyper-parameters. The best combination of hyper-parameters had the lowest 5-fold cross-validated mean reconstruction error and free energy. The best set of hyper-parameters was then used to train the model on the full dataset to obtain the final model and its parameters (W, a, b). The model was set to run for 1000 epochs, and the progress of the training was monitored for convergence using the cross entropy between the binary input and reconstructed visible variables,^30,31^ the mean squared deviation between the continuous input and reconstructed visible variables, and the pseudo-likelihood and the percentage weighted difference in free energy between the training data (80%) and validation data (20%), all of which had to stabilize.

### Simulation with the Model

Simulation with the RBM follows the block Gibbs sampling algorithm to sample from marginal distributions of interest by fixing some visible nodes so as to generate the remaining visible nodes to complete the observation. This was done by starting with an initial visible state made up of the original input data for the fixed variables corresponding to a partial observation and assigning random values for the variables we wanted to predict (termed missing variables in this case and does not refer to data “missingness”). This was followed by an alternate sampling of all hidden nodes with equation (3) followed by sampling of visible nodes with equation (4) or (9). However, after each Gibbs sampling block, the fixed variables were updated to those in the initial visible state while the missing variables were kept as those generated by the Gibbs chain. The Gibbs chain was continued until convergence to a stationary state of the visible nodes. For each individual and each row in the data, 100 simulated datasets were generated using 100 Gibbs chains. The simulation procedure was followed for the model evaluation step and for the model application for TDM.

### Model Evaluation and Generalizability

The goodness-of-fit evaluation was aimed at determining the level of agreement between the modeled and observed statistical distributions and utilized the categorical visual predictive check (VPC) and histograms of simulated proportions of PROMS. The categorical visual predictive check (VPC) plot is similar to figure 6 in Bisaso et al^9^ was generated by simulating 100 new datasets using the model as described in the previous section by fixing the independent variable (time) and covariates (mid-dose concentration, demographic and clinical) and the baseline PROMs after each full Gibbs iteration to the initial value while letting the dependent variables (the post-baseline PROMs) to follow the Gibbs chain sampled values up to stationarity. For each simulation, the proportion of each reconstructed PROM was plotted against time and superimposed with the observed proportion of that PROM to determine the level of agreement between the observed and simulated trends. The histograms of the simulated proportions of PROMs at each time point were generated and superimposed with proportions of the same PROMS to determine the agreement between the observed and simulated distributions.

The generalizability of the model was monitored during the process by subtracting the average free energy of a left-out dataset from the average free energy of the training dataset for each epoch. The model was taken to be generalizable when the difference in the training and validation datasets average free energies was close to zero. The model was deemed suitable for use if it passed both the goodness of fit and generalizability criteria.^32^

### Model Application

#### Variable Importance Ranking

Identification of the key variables associated with or predictive of the outcome variables is routinely undertaken in pharmacometrics to arrive at parsimonious, better fitting, more understandable, and implementable models that can help reduce the costs of future experiments.^33^ In line with that objective, the RBM model was used to rank the importance of mid-dose concentration, viral load, CD4 count, and baseline PROMS in relation to the post baseline PROMS.

A perturbation-based approach was used to determine the importance of variables in the trained model.^34^ The difference between the free energy of the original dataset and the free energy of the dataset with the *i*^*th*^ variable systematically perturbed (rearranged) was calculated for each individual record as

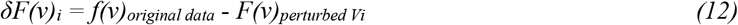

The variable with the larger mean increase in free energy was more important. Additionally, the Wilcoxon signed-rank test was used to test for the significance of the difference in free energy for each of the variables. Additionally, the mean changes in free energy were normalized to sum to one, and a barplot of variable importance was generated.

#### Target Concentration for TDM

For illustrative purposes, the model was used to generate the trend in NPIs when the mid-dose concentrations were capped at 4 mg/L as recommended for efavirenz TDM.^41^ To achieve this, the mid-dose concentrations in the original dataset that were greater than 4 mg/L were set to 4 mg/L, leaving the rest unchanged. The block Gibbs simulation approach was applied to the data to generate NPI symptoms on days 14 and 84. The results were plotted and superimposed with the simulations from the unaltered mid-dose concentration data.

## Results

The final model had 200 hidden units and 30 visible units corresponding to the input data. Figure 2 shows the structure of the model and the format of the training dataset.

**Figure 2.**
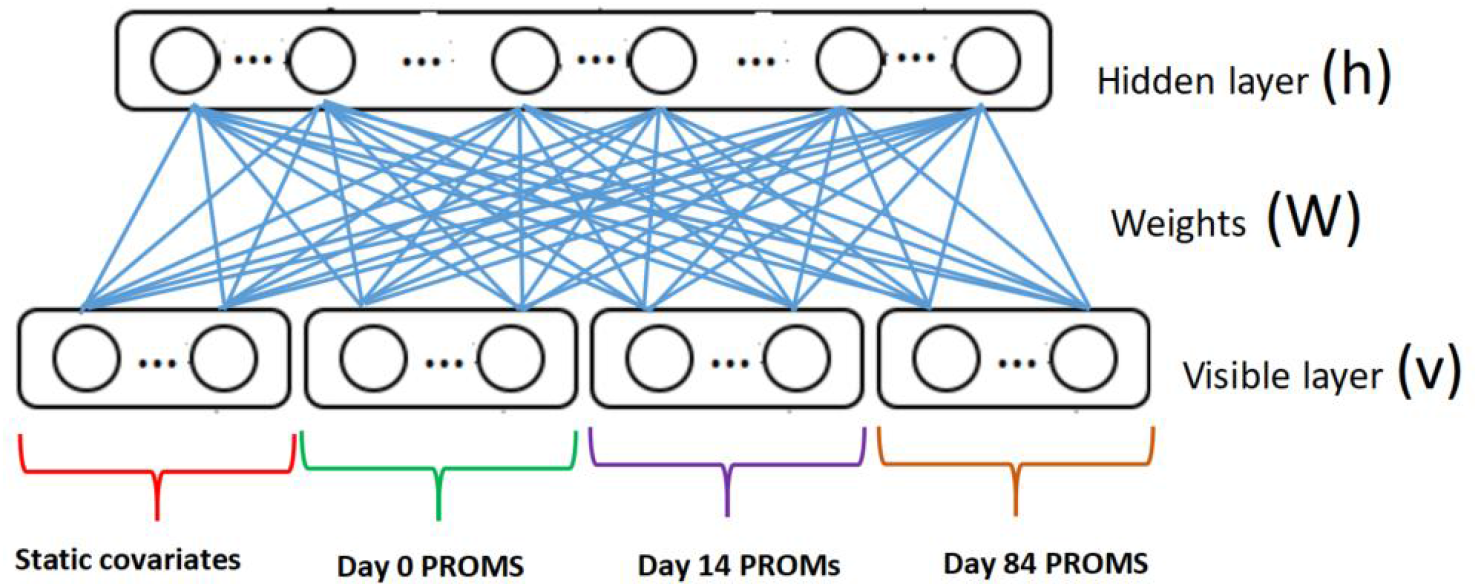
The architecture of the restricted Boltzmann machine model used to model the neuropsychological PROMs. The model was trained with a batch size of 30, a learning rate of 0.0001, a weight-decay of 0.0001, a momentum of 0.6, and no free-energy constraint on the cost function. The training process was achieved through the persistent contrastive divergence approach with 20 Gibbs chains and a 0.5 adversarial contribution to the gradient. The training process converged before the 1000 epochs elapsed (Figure 3).

**Figure 3.**
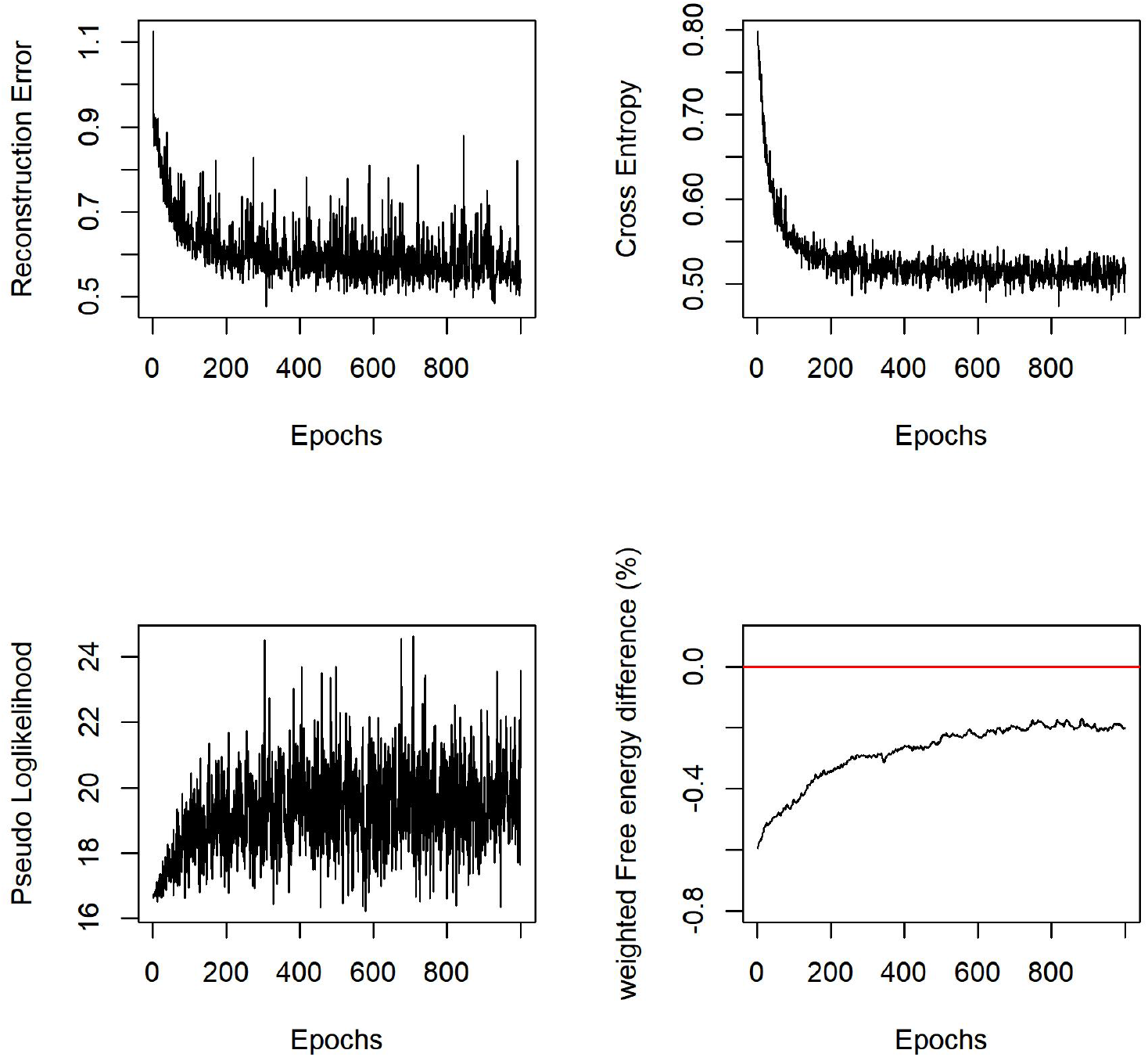
The trajectory of the training process of the final RBM model. The importance of the variables in the final model is shown in Figure 4. Noteworthy is the mid-dose concentration ranking lower than CD4 count and viral load relative to the post baseline PROMS. Baseline hallucinations (visual, audio, and tactile) showed the least change in free energy when perturbed.

**Figure 4.**
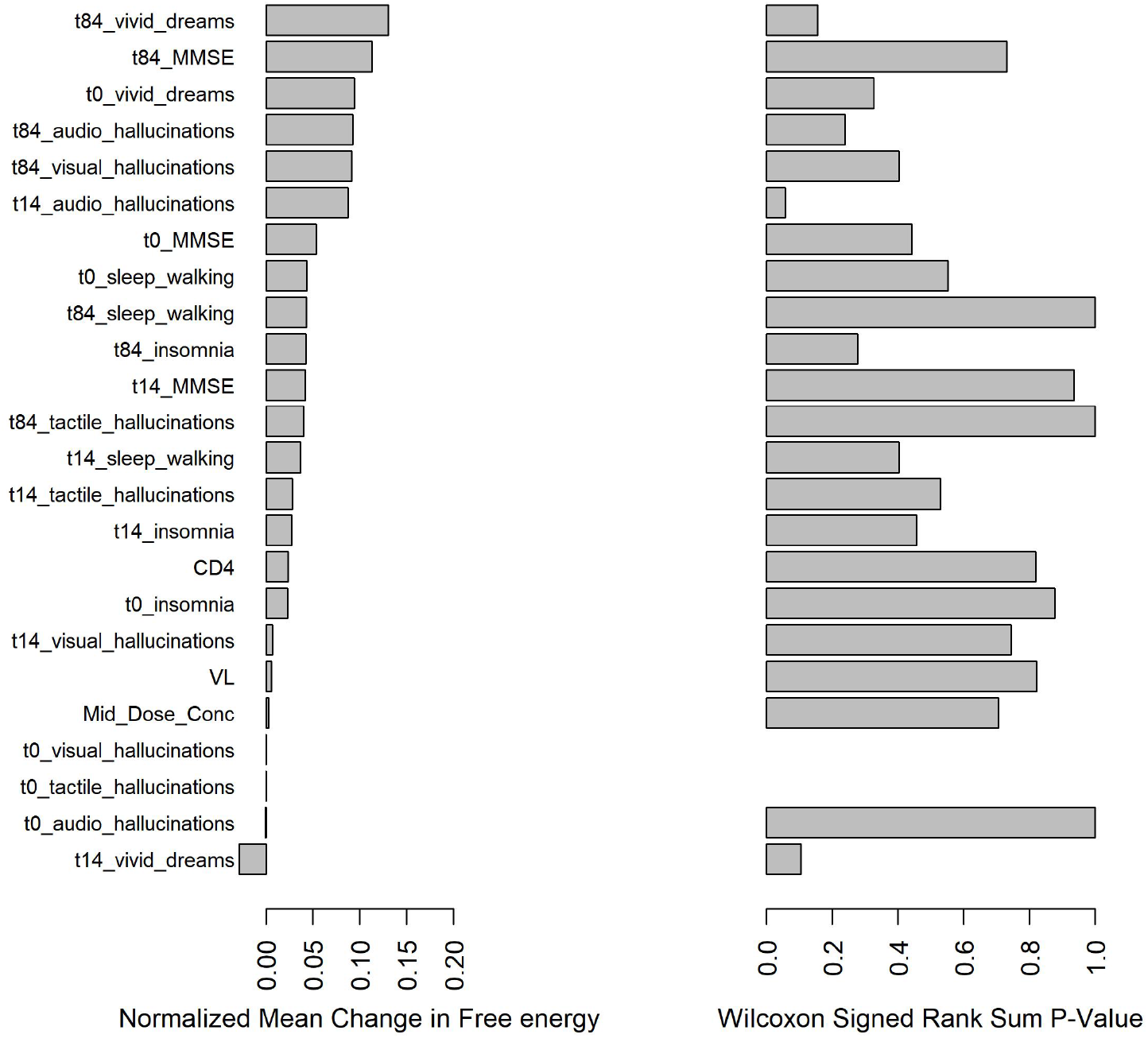
Ranking the importance of variables in the final model. t0_variable = variable at time 0,t14_variable=variable on day 14, t84_variable = variable on day 84. VL=Viral load, TB=TB disease status. As shown in the categorical VPC (Figure 5) and histograms of proportions of PROMS (Supplementary Figure S1), the model adequately learned the post-baseline trend of most PROMs.

## Discussion

Pharmacometric models serve three major objectives namely, to characterize the relationship between the primary independent and dependent variables, describe the effect of other variables (covariates) on that relationship between the primary variables, and to sample from the random effects statistical distributions to generate new data points that are statistically similar to the original data points via simulation. They are utilized for clinical trial design, efficacy comparison, dose optimization, and endpoint analysis.^35^ Pharmacometric models are typically nonlinear mixed effects models (NLMEM), that explicitly characterize the relationship between variables using a framework that comprises fixed effects and random effects sub-models. The fixed effects sub-model explicitly assumes and defines the primary trends in the relationship between observations using non-linear mathematical functions whereas its variation between individuals is described by the inter-individual (IIV) random effect sub-model.^36^ The unknown aspects of the modeled trend are captured by the residual random effect sub-model. Both the IIV and residual random effects sub-models assume a predefined statistical distribution.^37^ Incorrect or violated fixed and random effects model assumptions result in model misspecification and biased parameter estimation and prediction of newly simulated data points.^37–40^.

Generative RBMs are capable of achieving the three major objectives of pharmacometric modeling without making any assumptions regarding the functional nature of the trend/relationship between variables or their variability between individuals. They are particularly effective when dealing with multi-item PROM data, such as the NPI data used in this investigation, because of the complex relationship between the items as well as their joint relationship to clinical measures (viral load and CD4 count) and the underlying disease progression.

Furthermore, the variable importance ranking approach is based on the change in free energy, which is fundamental to how the RBM works.^17^ The energy function is directly related to the joint probability distribution and, therefore, captures complex relationships and interactions between variables.

Steady-state mid-dose concentration is used for the TDM of efavirenz.^41^ The generative stochastic modeling approach adapted for pharmacometric analysis of the data (i.e., pharmacometric generative stochastic modeling) revealed that baseline NPI symptoms and clinical variables (viral load and CD4 count) were more closely related, if not influential to the post-baseline NPI symptoms than steady state mid-dose concentration. This could be interpreted to mean that the observed trend in neuropsychological impairment is better predicted from baseline neuropsychological status than from mid-dose concentrations that are commonly used in TDM or baseline HIV severity. In previous analyses, we used the total exposure to drug (area under the concentration response curve)^7,9^ as the pharmacokinetic measure, while in this analysis, we use the mid-dose concentration. The previous analyses sought to establish the effect of dose reduction on the probability of NPI, while here we are interested in TDM. The current and previous findings could together be interpreted to mean that taking a one-off mid-dose concentration, as occurs in TDM, may not be as informative as determining the total exposure as far as predicting NPIs is concerned. The determination of what exposure metric, AUC or mid-dose concentration, to use in efavirenz TDM is outside the scope of this paper.

The model was used to simulate the proportions of NPIs when all mid-dose concentrations were capped at 4 mg/L, the conventional efavirenz therapeutic window’s upper bound. The resultant median trend in NPIs varied across individual items, with some showing only slight increase, others a slight decrease, or no change (figure 6). This non-uniform trend is also consistent with the variable importance result that showed mid-dose concentrations to be less influential on post-baseline NPIs than clinical status (viral load and CD4 count) and baseline NPI status, pointing to HIV disease as the principal cause of NPI symptoms. Noteworthy is the ability to generate individual patient trends, which is useful for the individualization of therapeutic and other interventions.

The RBMs are able to learn and approximate any function when given a large enough number of hidden nodes^42,43^ The downside to this is the tendency to learn the noise resulting in over-fitting the input dataset, limiting their prediction of unseen data. In this study, the optimal number of hidden nodes was selected as a hyper-parameter via 5-fold cross-validation on left-out data to maximize the RBM prediction on unseen data. Overfitting was also controlled by applying weight decay on all parameters during model training. Additionally, the weighted difference between the free energy of a training dataset and that of a left-out validation dataset was monitored throughout the training process. The percentage-weighted difference reduced and remained close to zero throughout the training process, indicating minimal overfitting.^19^

Graphical evaluation of the RBMs is simulation-based because of their generative nature that allows sampling of new data points from the learned distribution. This allows them to readily adapt to pharmacometric analysis, which utilizes simulation-based VPC for model validation. Indeed, the RBM fits the data better than the IRT model and was able to characterize more items of neuropsychological impairment, as can be seen from the VPC in Figure 5 compared to Figure 1 of Bisaso et al.^9^

**Figure 5.**
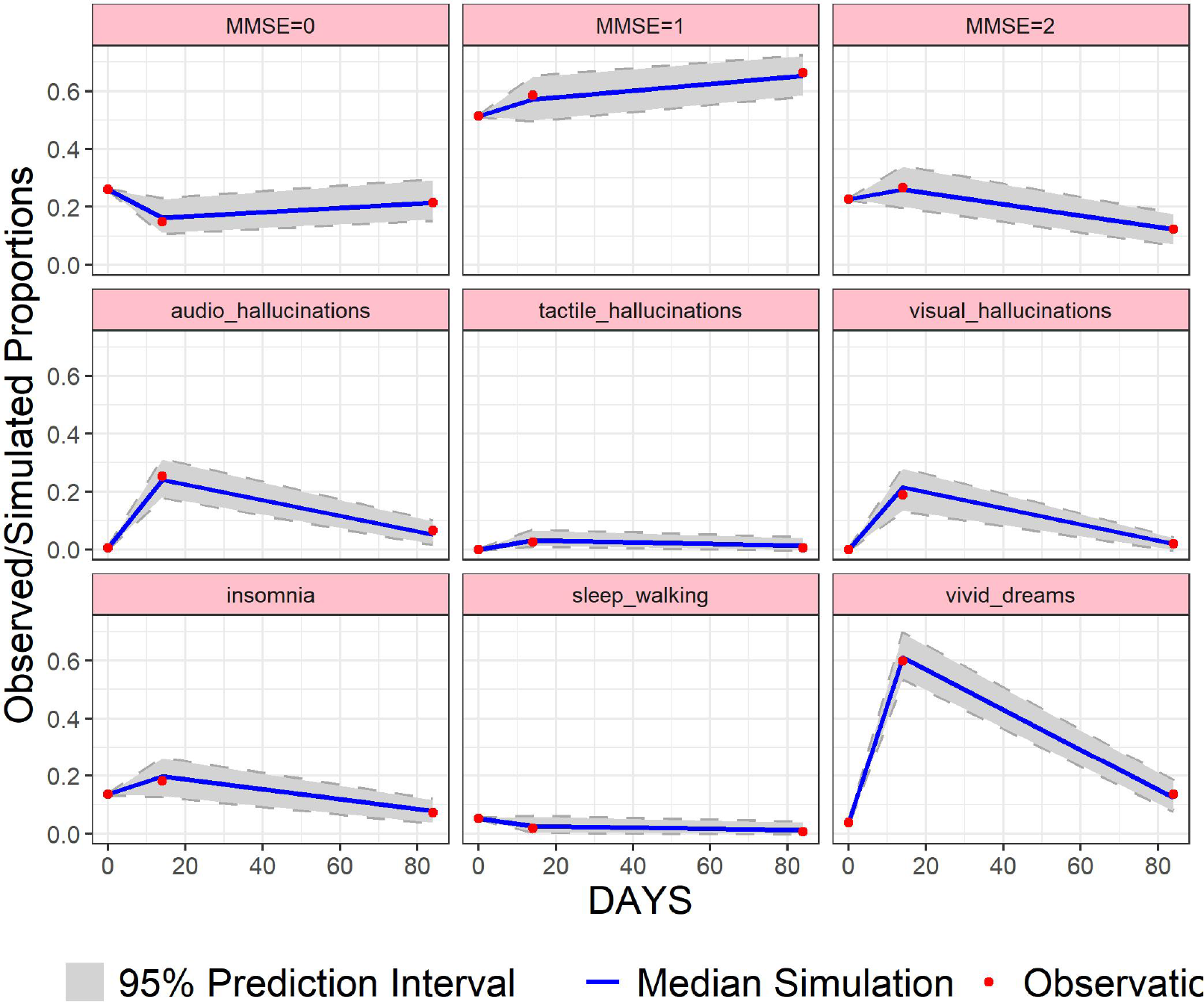
A visual predictive check depicting the model’s description of the time course of neuropsychiatric impairment symptoms. Solid black line. The simulation revealed an insignificant change in the trends of all NPIs with some reducing while others increased or remained similar (figure 6).

**Figure 6.**
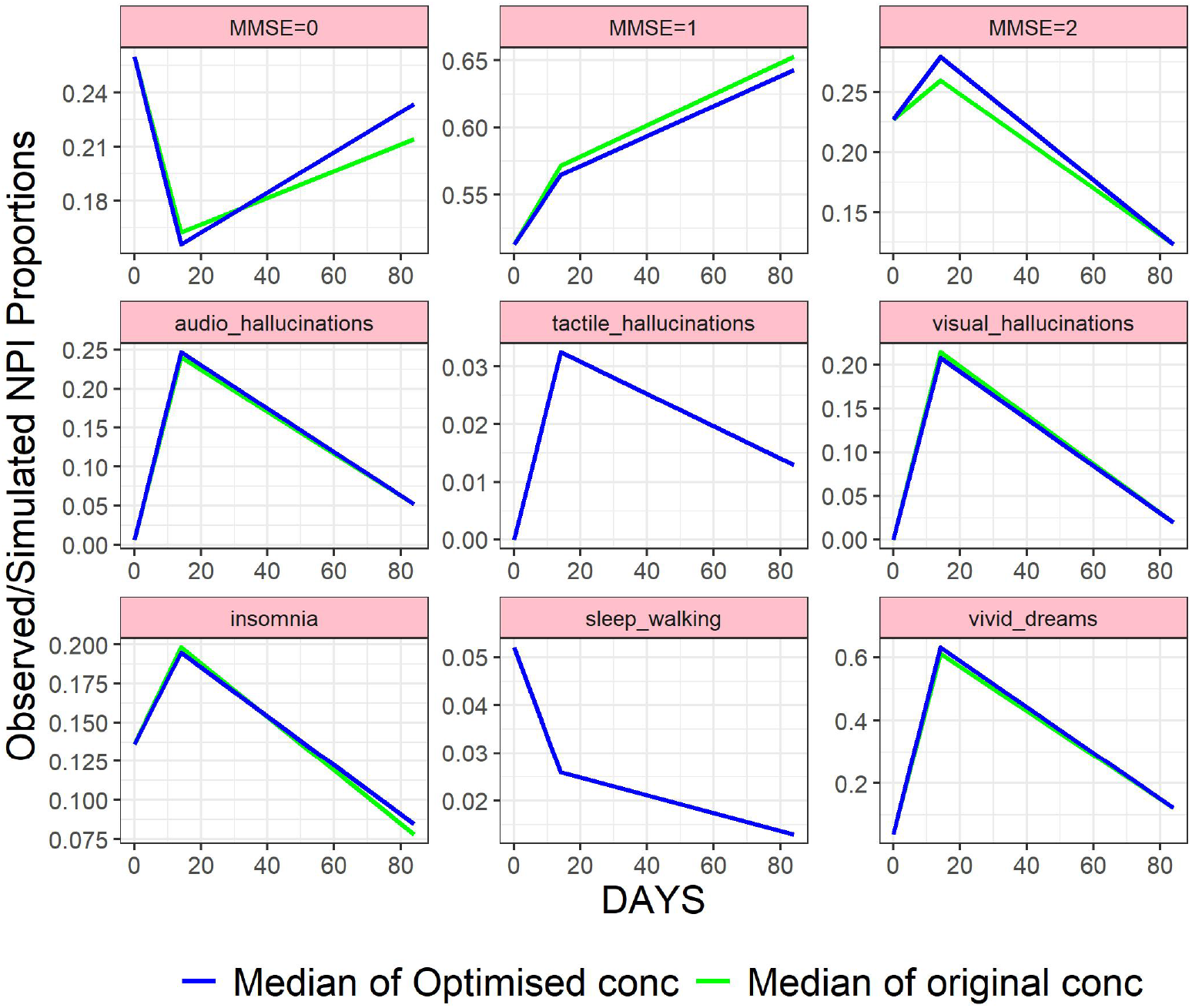
Predicted neuropsychological symptom proportions at both 400 mg and 600 mg doses of efavirenz and their corresponding 95% prediction interval.

Additionally, the model was able to generate the evolution of PROMs over time given the baseline PROMS, baseline clinical variables, and drug concentrations. Although the development of mechanistic models is desirable for improving our understanding of the underlying disease, as was done in Bisaso et al,^7,9^ the overwhelming complexity of multi-item PROMs makes it a daunting task. The ability of RBMs to directly infer the dependencies between multiple variables across time makes them attractive for modeling PRO disease progression. Moreover, in this study, there was no need to assume a pharmacodynamic functional relationship to neuropsychological impairment. Instead, the drug concentrations were directly used in the model with the PROMs indicating the simplicity of both modeling and model application for therapeutic drug monitoring or individualized target concentration intervention.

The RBM was chosen over other generative approaches because of its simplicity (one neural network with just 2 layers) and its grounding in statistical physics (laws of nature) as an energy-based model. Its simplicity eases training, modification, and application. Its physics grounding implies it has some underlying mechanism to it that tames its data greediness. Moreover, the intuition that low compatibility between variables is assigned higher energy and vice versa enables it to capture and represent complex multi-modal dependencies.^17^

Unlike traditional pharmacometrics or systems pharmacology modeling based on reductionism, generative stochastic modeling enables us to think holistically and integratively about the system under investigation. Although reductionism has been the bedrock of science investigations, pharmacology and pharmacometrics inclusive, we are still falling short in our understanding. The current approach to systems pharmacology is based on knowing the parts of the system with the intent that they add up to the whole. This may not always be the case, and the sum of the parts may not always be equal to the whole.^44,45^ There is, therefore, the need to apply systems thinking (i.e., holistic and integrative thinking) coupled with reductionism to the way we study drug action and response. This is where deep learning, in particular generative stochastic modeling, comes in. In this study, we have demonstrated how we use a systems thinking approach, through the use of generative stochastic modeling, to gain knowledge about neuropsychological symptoms in patients with HIV on efavirenz-based antiretroviral therapy, understand the importance of PROMS, pharmacokinetic and other variables in the modeled system without the need to understand the different mechanistic interactions.

## Conclusion

Restricted Boltzmann Machines are simple yet powerful models that are capable of accurately learning multi-domain patient-reported outcomes such as neuropsychological impairment in HIV patients and their evolution across time. They utilize the available phenotypic data without a need for mechanistic functional derivation or assumptions to learn the joint distribution from which new samples can be generated. They are capable of utilizing baseline data to generate individual-level trajectories of disease progression. They are readily adaptable to the common pharmacometrics workflow, thereby lending the benefits of assumption-free universal function approximation and generative power to model informed drug development with PROMs as outcome variables and use in TDM.

## Data Availability

All data produced in the present work are contained in the manuscript

